# Health System Determinants of Hypertension Care and Outcomes in Sub-Saharan Africa: A Systematic Review

**DOI:** 10.1101/2022.10.19.22280830

**Authors:** Samuel Byiringiro, Oluwabunmi Ogungbe, Yvonne Commodore-Mensah, Khadijat Adeleye, Fred Stephen Sarfo, Cheryl R. Himmelfarb

## Abstract

**Background:** Hypertension is a significant global health problem, particularly in Sub-Saharan Africa (SSA). Despite the effectiveness of medications and lifestyle interventions in reducing blood pressure, shortfalls across health systems continue to impede progress in achieving optimal hypertension control rates. The current review explores health system factors contributing to hypertension outcomes in SSA.

**Methods:** The World Health Organization health systems framework guided the literature search and discussion of findings. We searched PubMed, CINAHL, and Embase databases for studies published between January 2010 and June 2022 and followed the Preferred Reporting Items for Systematic Reviews and Meta-Analyses guidelines. We assessed studies for risk of bias using the tools from Joanna Briggs Institute.

**Results:** Thirty-nine studies clustered in 10 SSA countries met inclusion criteria. Health system determinants included human resource factors such as providers’ knowledge and adherence to hypertension treatment guidelines (n=21) and task sharing and shifting strategies (n=10). The second health system factors explored in service delivery were the health facility type and capacity (n=7) and hypertension service accessibility by cost, place, and time of services (n=15). A quarter of the included studies explored supply chain management for access to essential equipment and medicines. An additional set of studies addressed quality improvement strategies involving cross-integration of services (n=7) and various strategies of gauging the systems for better hypertension outcomes (n=8).

**Conclusion:** A combination of multiple rather than solo system interventions may yield significant improvements in blood pressure outcomes. Health information management and leadership involvement were less explored. Additional research on health system determinants of hypertension is needed to drive global improvements in hypertension outcomes. Future research would benefit from more rigorous implementation type interventional studies comprehensively assessing health system factors that contribute to better hypertension outcomes.

## Introduction

Hypertension remains a global health challenge, especially for low- and middle-income countries (LMICs) in Sub-Saharan Africa (SSA) [1]. Hypertension is a significant risk factor for stroke and ischemic heart disease, the world’s deadliest maladies [2]. It is estimated that 30% of the SSA population have hypertension [3–5], 27% are aware of their hypertensive status, and only 7% have their hypertension under control [3]. The prevalence of hypertension in SSA will likely continue to rise due to the rapid shift to sedentary lifestyles, growing urbanization, and a higher number of people living longer [6, 7]. The level of preparedness of health systems for managing hypertension and related sequelae is poor and requires urgent attention [8, 9].

Several strategies have been implemented to manage hypertension in SSA. Population-based interventions such as educational campaigns to improve dietary practices by reducing salt consumption have proven cost-effective in lowering blood pressure among hypertensive patients [10–13]. Alternative interventions implemented to reduce dietary salt include increasing the availability and affordability of healthy foods and reformulation of processed foods with the help of government policies [10]. Other strategies including technology and non-technology based reminders targeted the hypertensive patients to improve their adherence to the prescribed pharmacologic and non-pharmacologic treatments [13, 14]. Despite the numerous, effective interventions targeting the patients with hypertension, the health systems are not equipped to provide the best quality care, yet, the health system is the current epicenter in the population-based hypertension [8, 9].

Multiple guidelines exist for screening, diagnosing, and treating hypertension [2, 15, 16]. The Pan-African Society of Cardiology task force convened in 2014 laid the roadmap to achieving 25% blood pressure control in Africa by 2025[17]. In 2021, the World Heart Federation laid an updated roadmap that identified the major roadblocks in hypertension diagnosis and management and recommended solutions for reducing uncontrolled hypertension by 30% by 2030 [18]. The lack of local hypertension treatment guidelines, protocols, and policies on the procurement and distribution of anti-hypertensives are some of the multiple identified government and health system roadblocks to achieving hypertension control goals [17, 18]. These roadblocks likely result in the lack of community sensitization on hypertension and service inaccessibility through high-cost and centralized hypertension care [19, 20]. According to the Chronic Care Model, the health systems’ role in hypertension management should be to support patients for self-management at home, set up systems to ensure patients follow up after clinical encounters, and decentralize hypertension care closest to patients [20, 21]. Health systems should provide access to specialists if needed, ensure providers are trained and have access to the latest guidelines and decision support, and document care to improve the quality of care [20, 21].

The World Health Organization (WHO) health systems framework highlights six critical elements to target while working to improve the quality of health services. These are the service delivery; health workforce; access to equipment, consumables, and technology; information systems; financing; and leadership and governance. In 2013, the WHO developed the Service Availability and Readiness Assessment (SARA) to map the progress in the six elements of the health systems[22]. The SARA tool has been extensively utilized in maternal and reproductive health and on a minimal basis in cardiovascular health care [23, 24]. The evidence on the health system determinants of hypertension care and outcomes is limited. The current review synthesizes the literature on the health system factors of hypertension care in SSA and identifies the gaps and opportunities for improving the quality of hypertension care.

## Methods

### Search Strategy

Collaborating with a scientific library informationist, we defined the search terms for hypertension service availability and delivery guided by the WHO health systems’ framework [25]. We Identified Medical Subject Headings (MeSH) terms (“Health Facilities,” “Health Services Accessibility,” “Health Care Delivery”); and hypertension (“hypertension” or “elevated blood pressure” or “raised blood pressure” or anti-hypertensive) in SSA. We searched PubMed, Cumulated Index to Nursing and Allied Health Literature (CINAHL), and Embase. The detailed list of search terms is in Supplementary File 1 (S-File 1).

### Study selection and eligibility

The Preferred Reporting Items for Systematic Reviews and Meta-Analyses (PRISMA) [26] guided this systematic review (S-File 2). We imported articles from the literature search into Covidence, and three investigators (SB, KA, and OO) conducted the title and abstract screening, full-text review, and final extraction. Two investigators appraised each article independently at each step, and the third investigator resolved any conflicts. Occasionally, the investigators discussed with the supervising faculty (YCM and CRH) to resolve disputes.

In the article search and screening, we included studies (1) published between January 2010 – June 2022; (2) conducted in health facility settings (hospitals, clinics, community health facilities) in SSA; (3) with randomized controlled trials, quasi-experimental, cohort study, case-control study, cross-sectional study designs, and quality improvement projects; (5) which explored the association between healthcare system factors and hypertension awareness, treatment initiation and adherence, and control. The delineation of the search to 2010 was influenced by another systematic review conducted in 2013 exploring the influence of the health system on hypertension awareness, treatment, and control in LMICs [27].

We excluded studies (1) merely describing the prevalence of hypertension and associated patient-level factors, (2) which investigated the health system interventions without exploring association with any of the listed hypertension outcomes, and (3) which explored hypertension care interventions conducted solely in the community setting without describing collaboration with health facilities, or (3) examining patient-level behavioral change interventions for hypertension management. Additionally, we excluded gray literature (books, cases, etc.).

The investigators tracked the rationale for excluding any study during the entire process following the article search as reported in the PRISMA flowchart (Figure 1).

**Fig 1.**
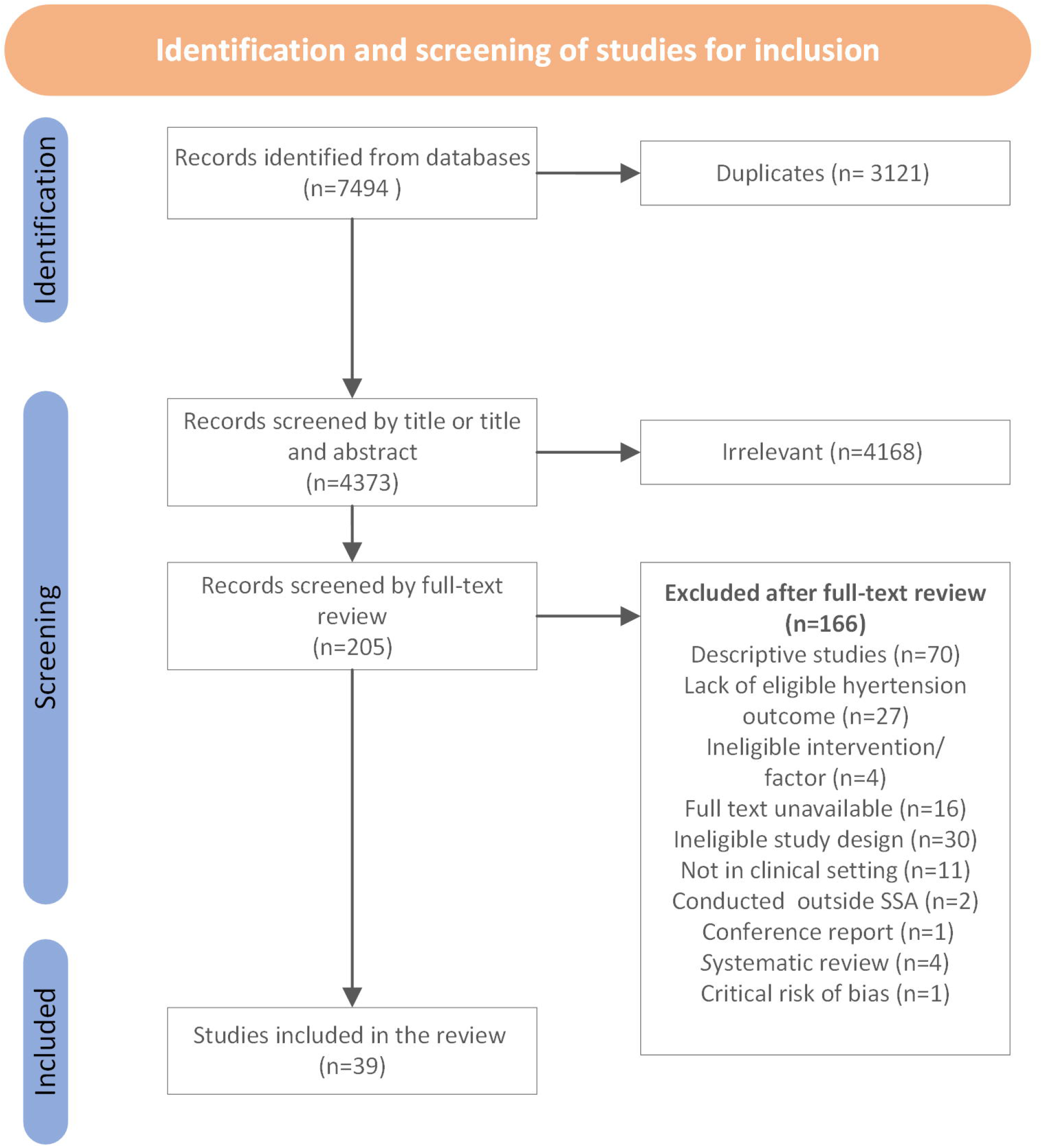
PRISMA diagram with flowchart of study screening and inclusion].

### Risk of Bias Assessment

We used the Joanna Briggs Institute (JBI) tools to assess the quality aspects of the methods and conduct of the studies included in the review [28]. We evaluated the quality improvement study using the Standards for Quality Improvement Reporting Excellence 2.0 (SQUIRES) guideline [29]. SQUIRES uses a checklist of standard elements for reports of system-level projects to improve healthcare quality. Two investigators independently assessed the risk of bias for each study and the third resolved conflicts. The JBI tool uses “Yes,” “No,” and “Unclear or Not applicable” to assess the different features of study design and the overall appraisal as include study, exclude study, or seek more information. We replaced “Yes,” “No,” and “Unclear” with “Low risk of bias,” high risk of bias,” and “Some concerns,” respectively. We assigned the overall quality appraisal of each study as “good,” “fair,” and “poor” and studies with poor quality were excluded. We used the robvis [30] online tool to create risk-of-bias plots where the overall appraisal was also converted to “low risk of bias” for the study of good quality, “some concerns” for studies of fair quality and “high risk of bias” for studies of poor quality.

### Analysis and synthesis of findings

After the data extraction, we identified relevant themes of health system factors and interventions for hypertension care. We classified the outcomes of each study in the appropriate group: hypertension awareness, systolic and diastolic blood pressure (BP), treatment initiation and adherence, and BP control. We tracked which factors/interventions are likely to lead to better hypertension outcomes and compared findings across the different countries of SSA. We discussed the results against the other potential areas of health systems crucial to improving the quality of hypertension care as delineated in the WHO health systems framework [25].

## Results

We identified 7,494 studies, yet 3,121 were duplicates (Figure 1). We conducted title and abstract screening for 4,373 studies yielding 205 for full-text review. The latter led to 39 studies for inclusion in this systematic review (Table 1). The quality assessment demonstrated a low risk of bias in 62% (24/39) of the included studies (S-File 3). Given the nature and complexity of health system interventions, the compliance to all requirements of the study design raised some concerns in 38% (15/39) of the included studies. Among randomized controlled trials, the most common issues were the lack of concealed intervention allocation, the similarity of groups at baseline, and blinding participants and those delivering the intervention of the group assignments. The most common concern among cohort studies was the lack of identification and management of confounding factors and strategies to address participants’ loss to follow-up. The identification and management of confounding factors was the most common reason for concern among cross-sectional studies.

**Table 1.**
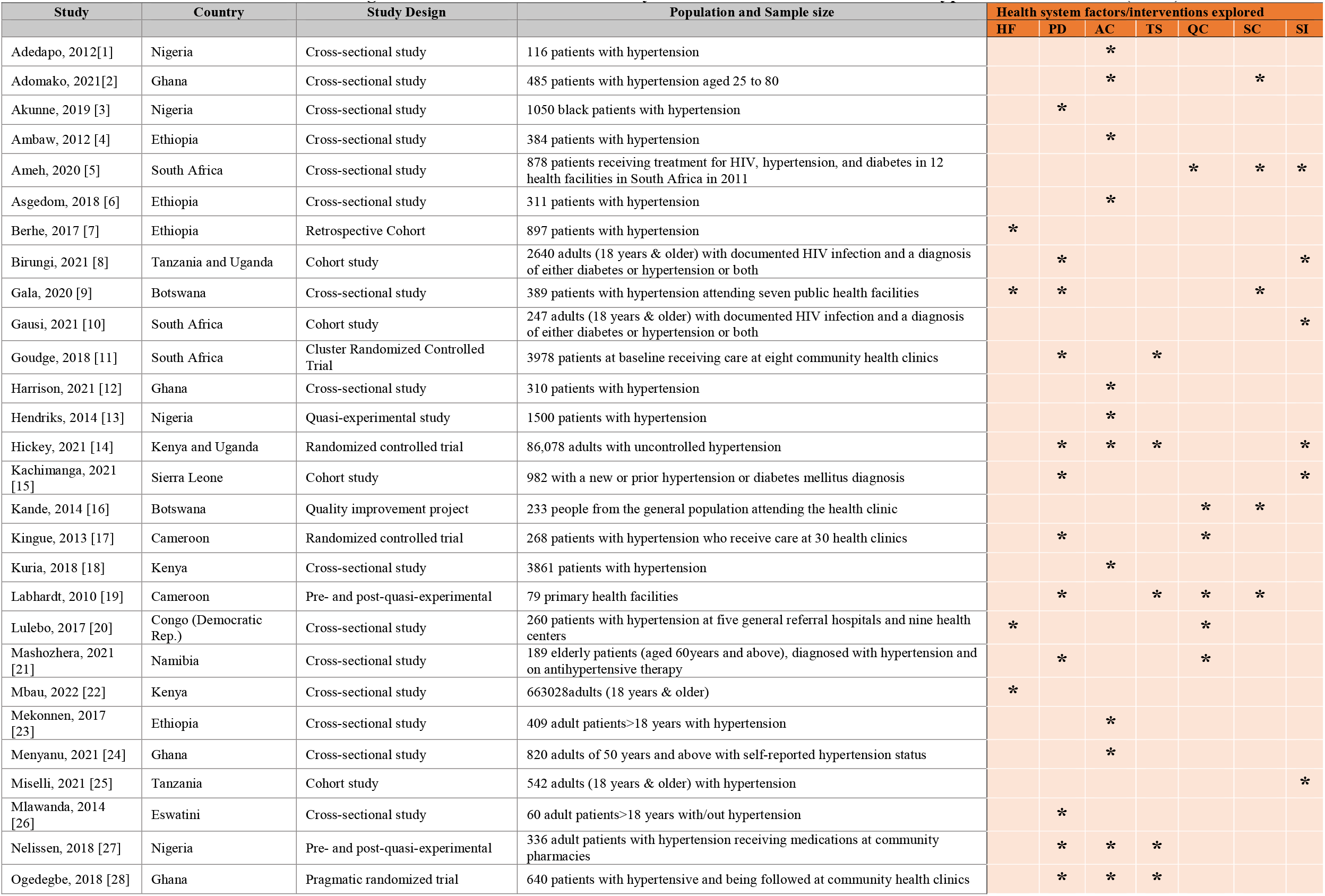
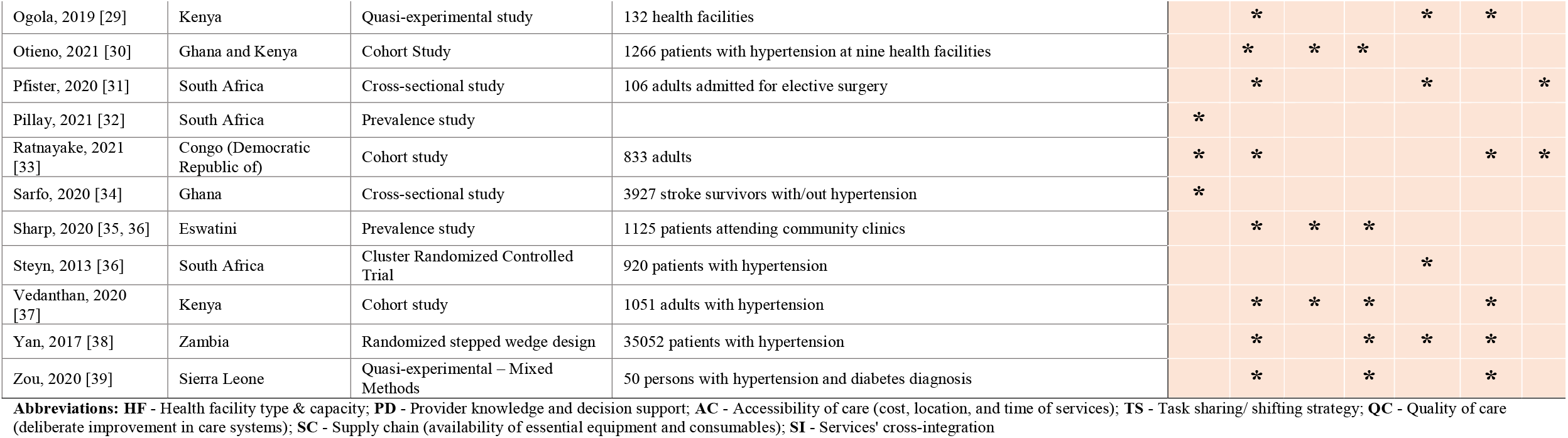
characteristics of studies examining the association between health system factors or interventions and hypertension outcomes (N=39)

### Country and design of included studies

Of the 39 included studies, six were randomized controlled trials conducted in South Africa (n=2) [31, 32], Cameroon (n=1) [33], Ghana (n=1) [34], Kenya and Uganda (n=1) [35], and Zambia (n=1) [36]. Five quasi-experimental studies were conducted in Nigeria (n=2)[37, 38], Cameroon (n=1) [39], Kenya (n=1) [40], and Sierra Leone (n=1) [41]. Eight cohort studies were conducted in Ethiopia (n=1) [42], Uganda and Tanzania (n=1) [43], Tanzania (n=1) [44], South Africa (n=1) [45], Sierra Leone (n=1) [46], Ghana and Kenya (n=1) [47], Kenya (n=1) [48], The Democratic Republic of the Congo (n=1) [49].

Nineteen cross-sectional studies were conducted in Ethiopia (n=3) [50–52], Nigeria (n=2) [53, 54], Botswana (n=1) [55], Democratic Republic of Congo (n=1) [56], Swaziland now called Eswatini (n=2) [57, 58], Namibia (1) [59], South Africa (n=3) [60–62], Ghana (n=4) [63–66], Kenya (n=2) [67, 68]. We additionally identified one quality improvement report from Botswana [69]. The geographic representation of included studies by country is demonstrated in Figure 2.

**Fig 2.**
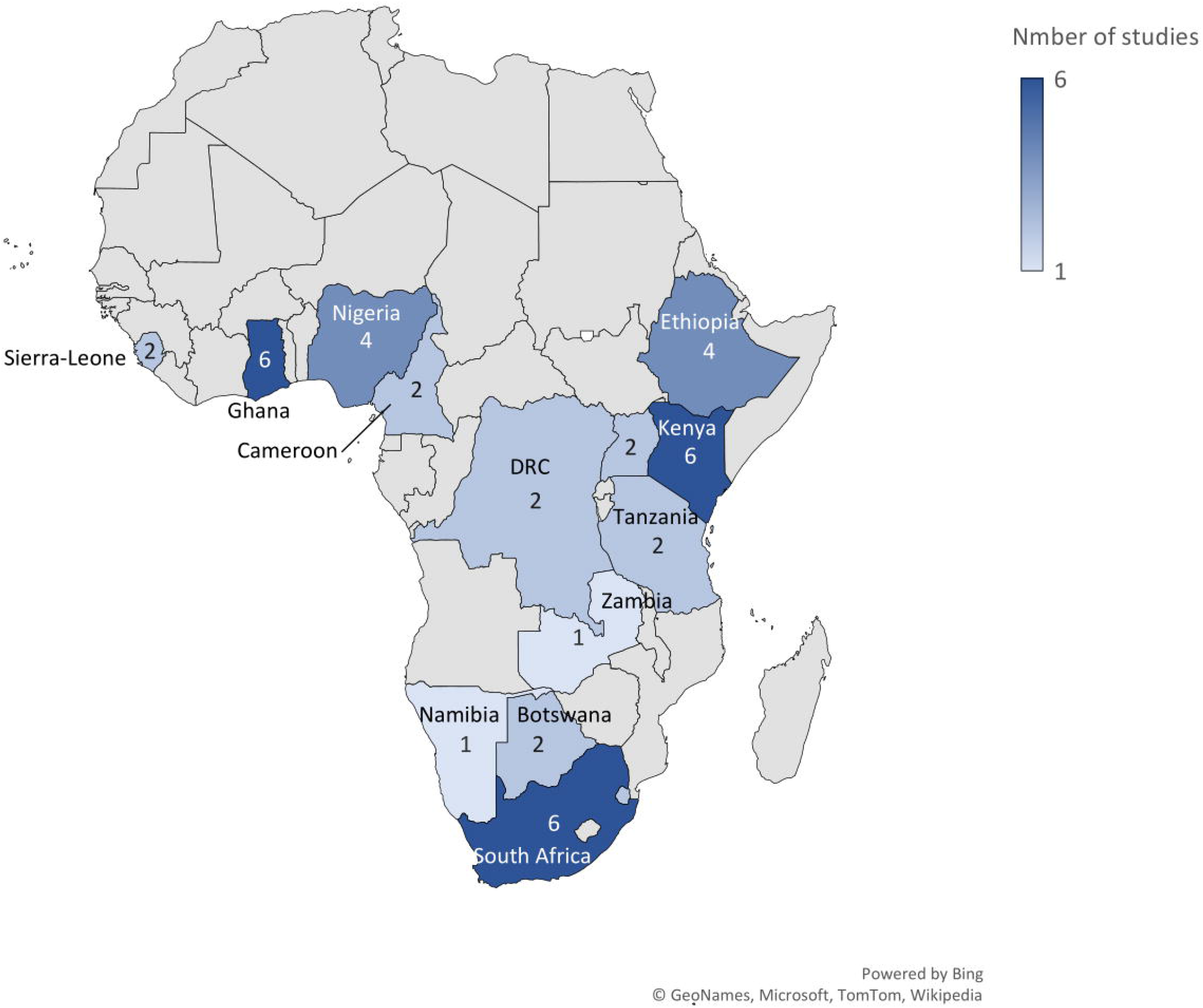
Distribution of studies on health system factors of hypertension care in Sub-Saharan Africa.

### Health system factors and interventions explored

The most studied health system factors were human resource factors – (1) healthcare providers’ knowledge and adherence to hypertension treatment guidelines and decision support systems; and (2) the task-sharing/shifting of hypertension care tasks to unconventional health professionals. Other health system factors related to service delivery included (3) the health facility type and capacity and (4) service accessibility in terms of cost, place, and time of care (Table 2). Additionally, studies explored (5) supply chain management for access to essential equipment and anti-hypertensive medicines. Finally, studies explored the revision of the patient care process at the health facility through (6) the cross-integration of services and (7) the different quality improvement strategies for improving hypertension care.

**Table 2.** Health system factors and interventions explored and corresponding countries where they were conducted.

| Health system factor/interventions | Number of studies | Country (number of studies) |
| --- | --- | --- |
| Health facility type and capacity | 7 | Ethiopia (1), Botswana (1), Democratic Republic of the Congo (2), Kenya (1), South Africa (1), Ghana (1) |
| Provider knowledge and decision-support | 20 | Nigeria (2), Tanzania (1), Uganda (2), Botswana (1), South Africa (2), Kenya (4), Sierra Leone (2), Cameroon (2), Namibia (1), Eswatini (1), Democratic Republic of the Congo (1), Zambia (1) |
| Accessibility of care (cost, location, and time of services) | 15 | Ethiopia (3), Ghana (4), Kenya (3), Nigeria (3), Eswatini (1), Kenya and Uganda (1) |
| Task-sharing/shifting strategy | 10 | South Africa (1), Kenya and Uganda (1), Cameroon (1), Nigeria (1), Ghana and Kenya (1), Ghana (1), Kenya (1), Eswatini (1), Zambia (1), Sierra Leone (1) |
| Quality of care (deliberate improvement of care systems) | 10 | South Africa (3), Botswana (1), Cameroon (2), Democratic Republic of the Congo (1), Namibia (1), Kenya (1), Zambia (1) |
| Supply chain (availability of essential equipment and consumables) | 10 | Ghana (1), South Africa (1), Botswana (2), Cameroon (1), Kenya (2), Democratic Republic of the Congo (1), Zambia (1), Sierra Leone (1) |
| Services cross-integration | 8 | South Africa (3), Tanzania and Uganda (1), Kenya and Uganda (1), Sierra Leone (1), Tanzania (1), Democratic Republic of the Congo (1) |

Note: Some studies reported more than one system factor

#### (1) Healthcare providers’ knowledge and adherence to hypertension treatment guidelines and decision support system

Half of the studies (21/39) [32–36, 38–41, 43, 46–49, 53, 55, 57–59, 61] across 13 countries explored the impact of healthcare provider knowledge of or adherence to hypertension treatment guidelines and decision support system on hypertension outcomes (Table 3). Generally, higher provider knowledge of or better adherence to guidelines and availability of decision-making support systems correlated with better hypertension care outcomes. For instance, the lack of physician knowledge in prescribing anti-hypertensives at a University College Hospital in Nigeria demonstrated 76% higher or lower blood level doses (as measured in urine) of anti-hypertensives than recommended and contributed to 35% lower BP control [53]. Adherence to the Eighth Joint National Committee guidelines contributed to 46% of additional patients achieving BP control in Kenya [70]. An approximately similar pattern was observed in a study conducted in Botswana [55].

**Table 3.**
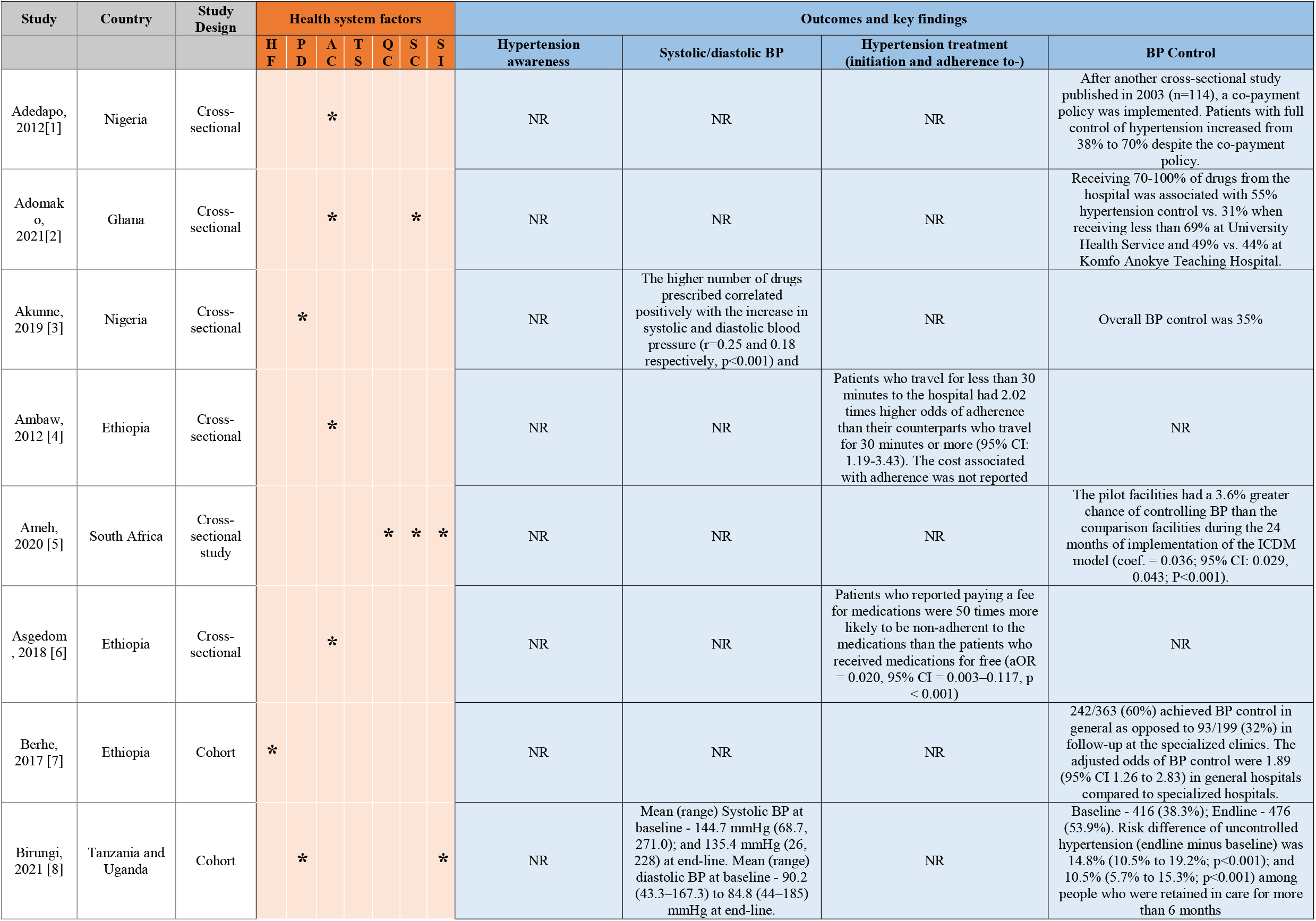
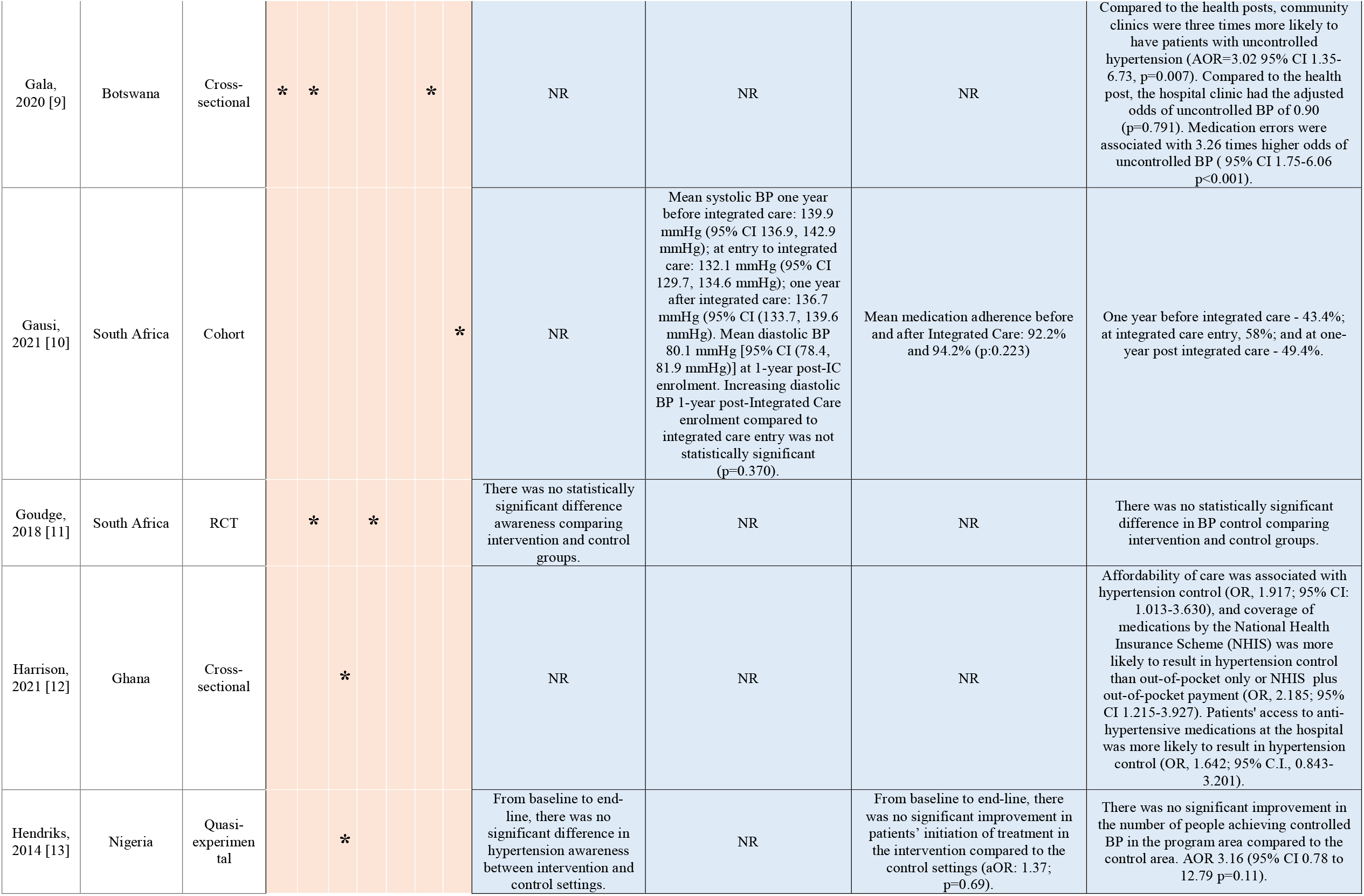
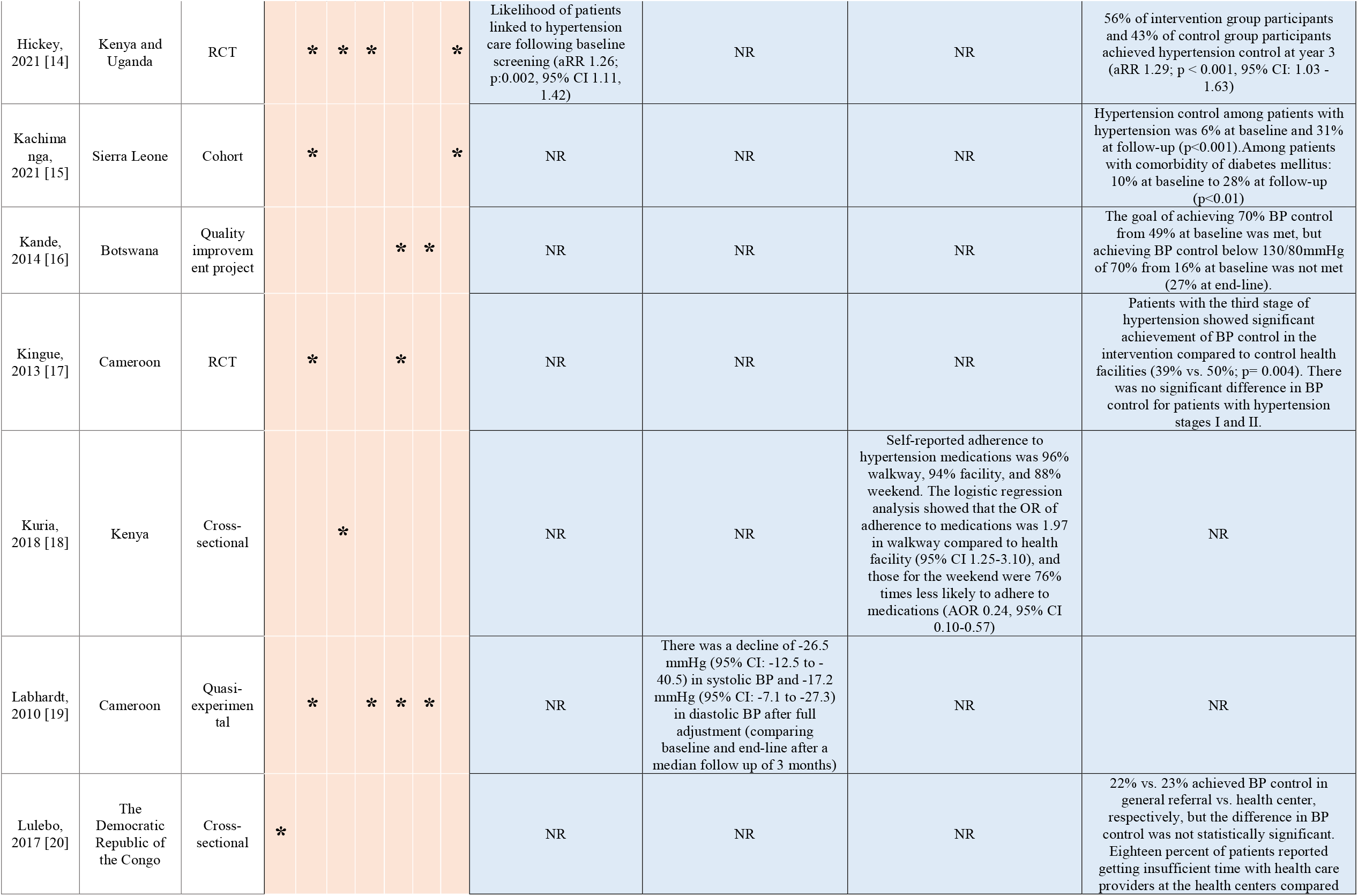
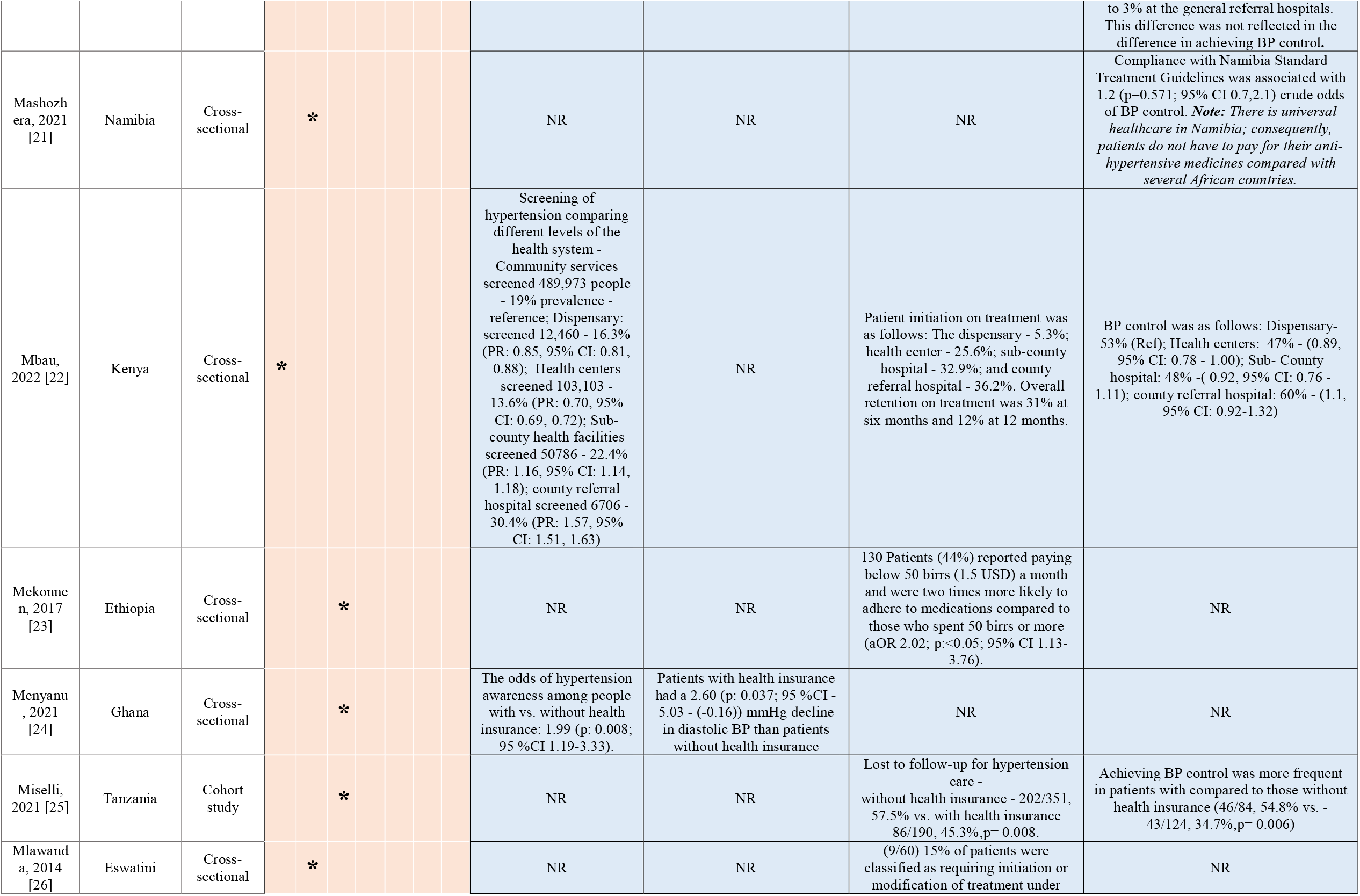
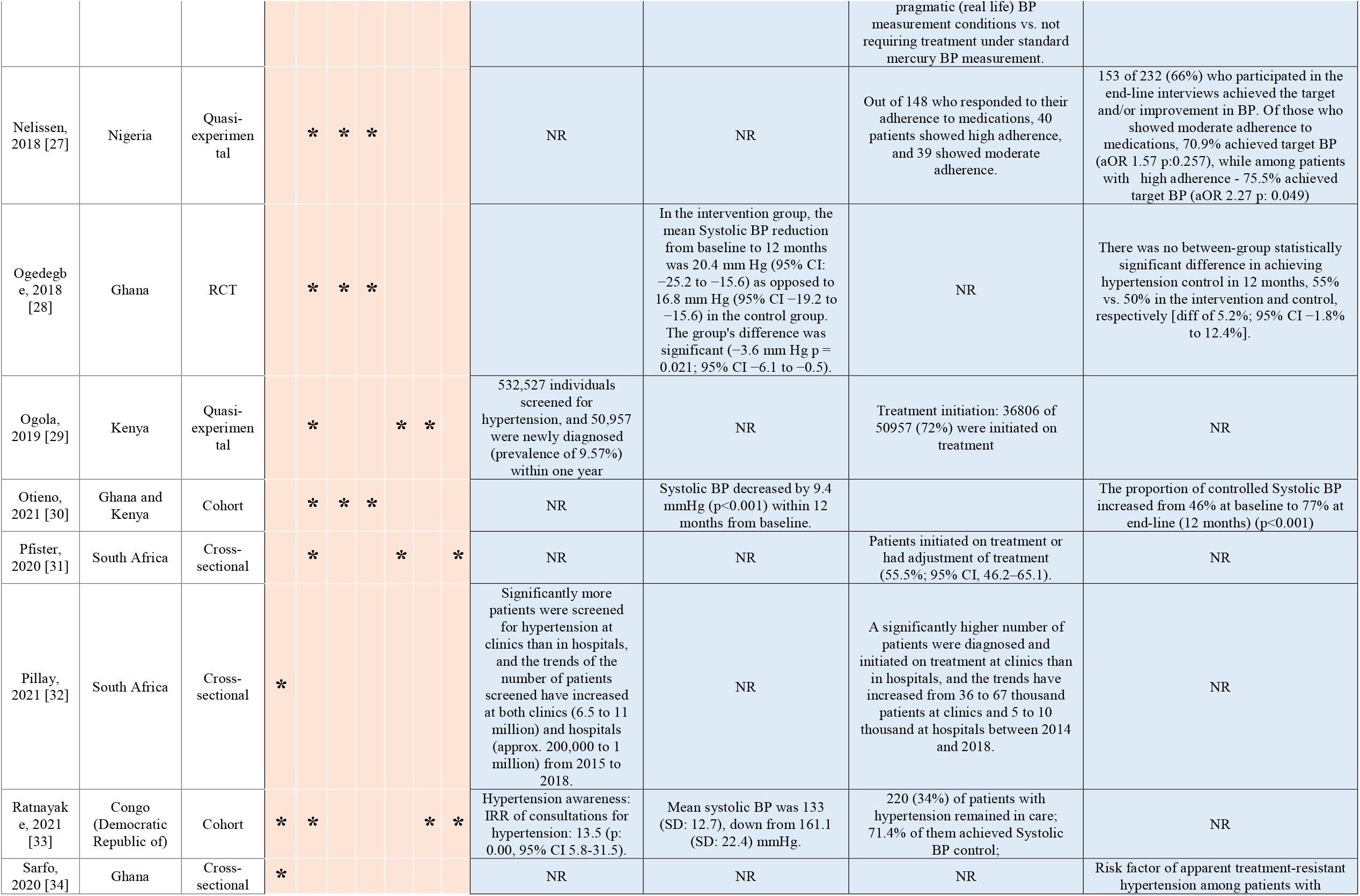
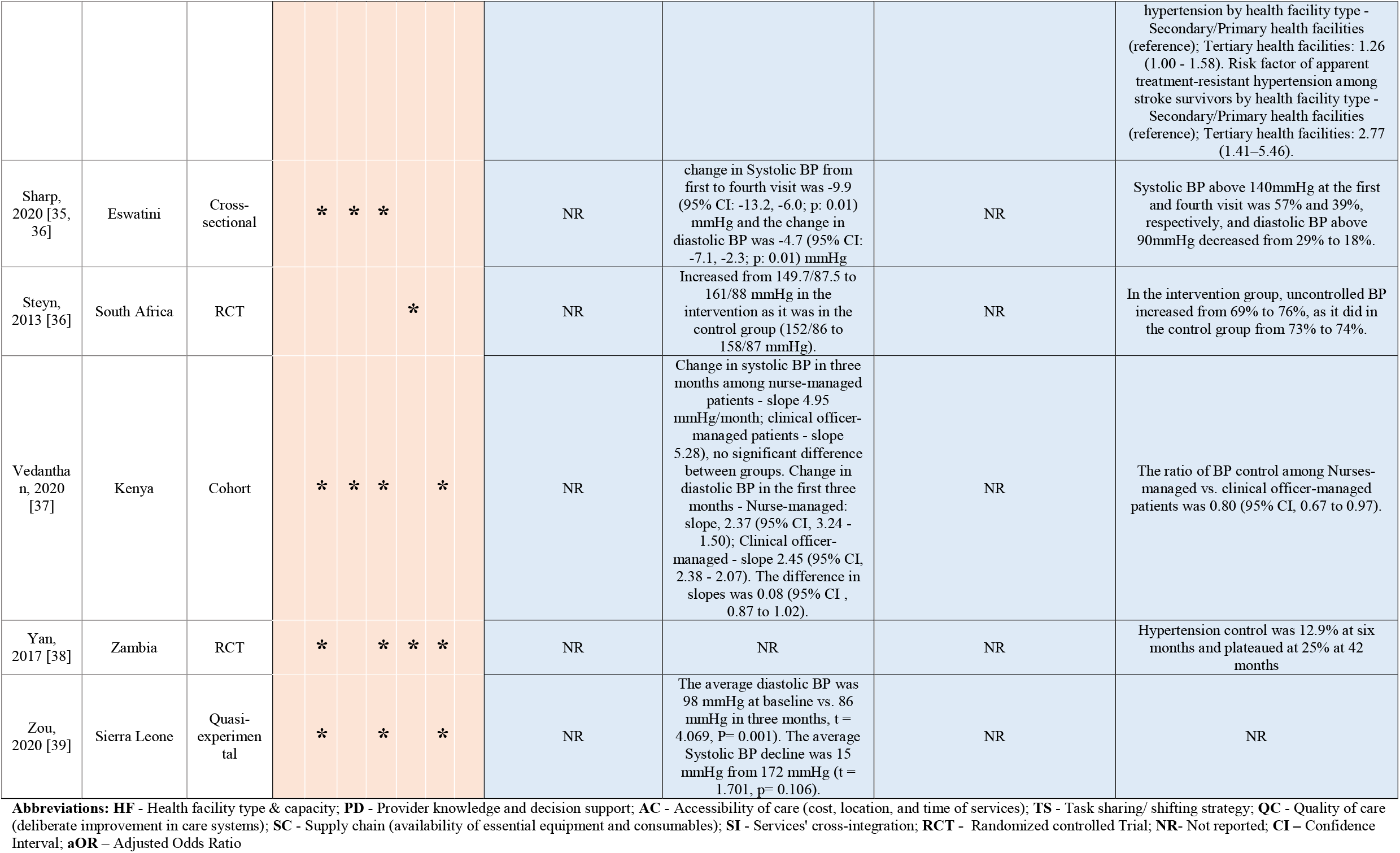
Studies examining the association between health system factors or interventions and hypertension outcomes (N=39)

Improvement in the healthcare providers’ skills and knowledge of the correct blood pressure measurement techniques and hypertension diagnosis, respectively, were effective in managing hypertension in Kenya [40]. In its first year, the program helped screen 532 527 individuals, diagnose almost 10% of them with hypertension, and initiate 72% of those diagnosed on treatment [40]. The provider skills in BP measurement were assessed in Eswatini by comparing the pragmatic measurements with standard sphygmomanometer measurements [57]. The two measurements showed a good agreement of 0.7 Kappa score and an 84% concurrence in hypertension treatment decisions (continuing, changing, adjusting, stopping treatment).

#### (2) Task-sharing/shifting strategy

A quarter of studies (10/39) [32, 34–36, 38, 39, 41, 47, 48, 58] conducted in 9 countries explored the impact of delegating some hypertension care tasks to non-physician providers (task-shifting) or community health workers. The task-shifting consisted of training and giving roles to lay community health workers in South Africa [32] and Zambia [36] to conduct and record the BP measurements, to nurses in Cameroon [39] and Ghana [34] to treat hypertension (including prescribing drugs), and to pharmacists in Nigeria [38] to treat hypertension (prescribe medications) and dispense medications after diagnosis at the health facility.

The impact of task-shifting varied across studies but bent towards positive BP outcomes. In Ghana [34] and Cameroon [39], task-shifting contributed to the significant decline in systolic and diastolic BP but not in the achievement of controlled BP [34, 39]. On the other hand, in the two quasi-experimental studies conducted in Nigeria and Zambia, respectively, the task-shifting contributed to the achievement of controlled BP in Nigeria (AOR 2.27, p: 0.049) from baseline to end line and in Zambia from 13% at baseline to 25% (no p-value reported) among patients who attended at least 2 follow up visits [36, 38]. In a cluster randomized controlled trial conducted in South Africa, the task-shifting to lay community health workers in BP measurement and documentation contributed to higher adherence to appointments (75% vs. 56% patients attending their assigned appointments) but no statistically significant improvements in achieving BP control comparing the intervention to the control group [32].

#### (3) Health facility type and capacity

Eighteen percent (7/39) of the included studies [42, 49, 55, 56, 62, 65, 68] explored the impact of health facility type and capacity on hypertension care and outcomes. The studies highlighted the significant yet inconsistent differences in hypertension outcomes associated with receiving care at either community health post (closer to the community - offering one or very few primary healthcare services), community health center (serving a slightly bigger size of the community - offers the whole package of primary care services), general referral hospital, or specialized clinics. In Botswana, community health posts were three times better than community health clinics in helping patients achieve BP control [55]. In Ethiopia, general referral clinics were almost two times better than specialized clinics [42] in patients’ achievement of BP control. In the Democratic Republic of Congo, there were no significant differences in BP outcomes between general referral hospitals and community health centers [56]. Additionally, the presence of the doctor on a daily basis (OR:1.64; 95% CI:1.17-2.31; *p*=0.004) and a higher mean number of nurses in the clinic (OR: 1.15; 95% CI:1.08-1.23; *p*=0.001) were significantly associated with BP control at the baseline of a randomized controlled trial conducted in South Africa [71].

#### (4) Accessibility of hypertension care (cost, geographic, temporality of service delivery)

The effect of patients’ access to hypertension care on hypertension outcomes was explored in 38% (15/39) of the included studies within seven SSA countries [34, 35, 37, 38, 47, 48, 50–52, 54, 58, 63, 64, 66, 67]. The general trend was that the lower cost of care was significantly associated with better hypertension outcomes. The two quasi-experimental studies conducted in Nigeria and Ghana provided health insurance coverage to the intervention group and to both study arms, respectively [34, 37]. In both studies, the systolic and diastolic BP declined in both study arms with a significant and sharper decline in the intervention arms even though the achievement of BP control was not significantly different in the intervention versus control arms. Two cross-sectional studies in Ethiopia reported that patients’ payment of user fees was associated with 50 times higher odds of uncontrolled BP while paying less than 50 Ethiopian Birr (approximately 1 USD) was associated with two-fold higher odds of achieving BP control [50, 52].

Four studies reported the factors related to the spatial and temporality accessibility of hypertension care services in achieving BP control. The shorter distance to the health facilities [51] and time flexibility at the hypertension care clinics were associated with better hypertension care outcomes [67]. Yet, such patterns were not true in South Africa, where rural dwellers (where people are likely to travel longer to health facilities) were better responders to treatment intensification by achieving BP control compared to urban dwellers (OR: 0.66; 95% CI: 0.45-0.97; p:0.034) [71]. In Ethiopia, traveling less than 30 minutes to the hospital had 2.02 times higher odds of medication adherence compared to traveling 30 minutes or more [51]. Similarly, the flexibility in location (other than at the health facility) and time of day of dispensing medications were associated with better health outcomes, as reported in Kenya [67] and Nigeria. [38] In Kenya, the extension of hypertension service hours to include evening hours led to a two times likelihood of patients’ adherence to anti-hypertensive medications even though patients scheduled to attend weekend clinics had 76% fewer odds of medication adherence [67].

#### (5) Supply chain management for the availability of essential equipment and consumables

A quarter (10/39) of the studies [36, 39–41, 48, 49, 55, 66, 67, 69] across eight countries explored the availability of equipment and consumables involved in hypertension management and impact on BP outcomes. The procurement of drugs and consumables and improvement in the supply chain implemented in the experimental studies and quality improvement project was in combination with other interventions such as providers’ training in Cameroon, Kenya, and Zambia [36, 39, 40], and better data management in Botswana [69]. In these studies, the impact of the equipment and supply chain was not necessarily isolated but undoubtedly contributed to the overall improvements in BP outcomes.

The cross-sectional study conducted in Botswana reported that the medication errors contributed to three times higher odds (Odds Ratio: 3.23; 95% CI 1.75-6.06 p<0.001) of uncontrolled BP. Almost one-third of the errors were dispenser errors caused mainly by the stock out of the medication and reluctance to refer the patients back to the prescriber to provide an alternative medicine [55]. The interventions that showed significant improvement in BP improved not only the availability of the equipment but also the essential anti-hypertensive drugs, as is the case in Kenya [40].

#### (6) Cross-integration of services

The integration of hypertension and other non-communicable diseases services into the well-established health services was explored in 18% (7/39) of the studies in 6 different countries [35, 43, 45, 46, 49, 60, 61]. The most frequent form of cross-integration was the provision of non-communicable diseases services in the Human Immunodeficiency Virus (HIV) and Sexually Transmitted Diseases clinics and the use of existing patient follow-up mechanisms in managing non-communicable diseases.

Predominantly, the cross-integration of services has shown significant improvements in hypertension control. A cohort of patients in South Africa increased in controlled BP from 43% before integration of services to 49% one year after integration [45]. Another cohort of patients in Uganda and Tanzania had a 10% decline in the risk of uncontrolled BP [43]. In Uganda and Kenya, the care for hypertension, diabetes, and HIV were integrated, and additional features of care improved, leading to a 26% higher likelihood of initiating patients on hypertension treatment and a 29% higher likelihood of BP control. In the Democratic Republic of the Congo, hypertension services were integrated into Emergency Primary Clinic. While there was a high rate of new hypertension diagnoses in 97% of the included sample, only 34% remained in care in one year, and other those who remained in care, 71% achieved a systolic BP below 140 mmHg, which is the cut-off used in the country for desired systolic BP [49].

In one instance, the hypertension clinic was built from scratch in Sierra Leone [46]. A cohort of patients followed in that new rural hypertension clinic had BP control go from 6% at baseline to 31% at one year-follow-up [46].

#### (7) Deliberate improvement in the quality of hypertension care

Eight studies [31, 33, 36, 39, 40, 60, 61, 69] conducted in 5 countries explored the additional value of deliberately gauging and improving systems for a better quality of hypertension care. The quality improvement projects included an improved data recording process in Zambia and Botswana [36, 69] and better monitoring through supervision in Cameroon [33] and South Africa [31]. In South Africa, a quality improvement project on improving data recording and adherence to treatment guidelines by providers was implemented through the introduction of a folded guidelines sheet inside the patient file [31]. The project did not contribute to an increase in patients with BP control, and providers reported that the project was overwhelming because of staff shortage [31]. Where quality improvement was implemented jointly with other system components such as improving the supply chain of equipment and consumables in Botswana [69]; providers’ training on hypertension care and task-shifting in Cameroon [33, 39]; supply chain, training, and task-shifting in Zambia [36]; there were better blood pressure outcomes.

## Discussion

This systematic review aimed to assess the health system factors and interventions for hypertension awareness, initiation and adherence to hypertension treatment, and achievement of controlled blood pressure in SSA. A 2013 systematic review in LMICs [27] and a 2015 narrative synthesis in SSA [72] explored the health system determinants of hypertension care and reported consistent findings of factors of hypertension outcomes: cost of care, human resources factors such as adherence to protocols, physical resources factors such as lack of functioning equipment, and lack of treatment guidelines. Our review identified the above and additional health system interventions and factors, including the accessibility factors of care such as place and time of care, the health facility type and capacity, the use of quality improvement strategies, and the cross integration of services.

The healthcare provider’s knowledge and adherence to hypertension treatment guidelines stood out as one of the most studied health system interventions [40, 73, 74]. The capacity building among healthcare providers and the task sharing and shifting of hypertension management roles to vital professionals like nurses, pharmacists, and community health workers align with scientific evidence from developing and developed countries [75, 76]. For instance, the nurses’ role in managing hypertension has evolved from merely measuring and monitoring blood pressure to collaboratively detecting, diagnosing, and referring patients with hypertension and its complications [77, 78]. In many countries and settings, nurses prescribe or dispense anti-hypertensive medications, provide patient education and counseling to ensure medication adherence, and assume leadership roles in spearheading quality improvement projects for better management of hypertension [77]. An estimation of the global gap in clinic visits for hypertension care reported that 50% of LMICs and 86% of lower-income countries have a physician deficit even if patients were to make only three annual visits to the health facilities for their hypertension [79]. Neupane and colleagues recommend shifting some hypertension management tasks to non-physician clinicians to bridge that gap [79]. Interdisciplinary collaboration is the way forward for SSA to provide quality hypertension care to patients; however, regional and national policies must align with and support best practices.

The cross integration of hypertension services is another system-level intervention explored for the management of hypertension. The hypertension services have been frequently integrated with HIV clinics, and this strategy has demonstrated to be effective in helping patients achieve controlled blood pressure in Sierra Leone [46], South Africa [45], and Tanzania and Uganda [43], to name a few examples. There are some reasons why taking lessons from the management of chronic infectious diseases could be effective in hypertension care. The management of chronic infectious diseases, HIV and Tuberculosis, in resource-constrained settings has, despite still-existing challenges, produced strong evidence and effective strategies for managing patients with such complicated chronic health conditions amid, just to name a few; insufficient healthcare professionals [80], inaccessibility of health services [81], and poor information management systems [82]. In addition to demonstrating that task sharing with and task shifting to non-physician healthcare providers is possible and effective [80, 83, 84], the process of managing HIV and Tuberculosis has also demonstrated ways to manage loss to follow up by the use of strong information management systems [82], decentralization of services close to the community [85], and collaboration with community health workers [80, 84]; all the strategies that could be well adapted to hypertension management. One of the information management system strategies that have been effective in HIV and Tuberculosis control and hypertension management in other low-income countries is the patient registries [86–89]. The current review did not find studies exploring patient registries to prevent loss to follow-up in hypertension management.

The hypertension clinics model consists of managing patients with hypertension in their own outpatient unit and was experimented and proved effective in Sierra-Leone [46] and Eswatini [58]. The premise of this strategy is to have well trained healthcare providers and good systems of data management, in some instances to decentralize of hypertension care, and improvement in patient follow-up. The lessons from the management of HIV and Tuberculosis could be applied in those services. It is crucial to ensure that the detachment of hypertension care from the general outpatient department or from the well-equipped hospital does not hamper the patients’ initiation on hypertension treatment through the internal transfer processes and that patients with hypertension continue to receive holistic care for other health problems they might have in addition to hypertension.

While the decentralization of care might address geographic barriers to hypertension care access, other factors of accessibility such as cost of care and time of services require additional and potentially different interventions. Extensively, the current and previous literature have shown that the high direct and indirect costs of care hamper patients’ use of health services and lead to poor health outcomes and among patients with hypertension, poor blood pressure control [54, 66, 90]. The time of operation of clinics, though limited evidence exists, has an impact on the staffing hence the long queues of patients at the in- and outpatient departments which discourages patients’ attendance to their appointments and encourages loss to follow-up [67, 91, 92]. Universal health coverage through government-subsidized insurance premiums has been advocated as a solution to prevent catastrophic spending on healthcare services and associated poor health outcomes [93–95], and the 24/7 hypertension clinic operation could be explored as a potential intervention to the long waiting queues at the clinics. The financing of universal health coverage and the 24/7 open clinics, however, require political buy-in and collaboration across sectors, local and international [94].

While leadership and governance are central to the success of any endeavor to improve the quality of care, their role and impact in advancing hypertension care in SSA are not widely researched, and the few existing studies are predominantly of poor quality [96]. Thornton highlighted the critical values of leadership commitment, willingness to fund care, strategic and creative local and foreign partnerships, and evidence-based guidelines as the key reasons why Botswana surpassed the United Nations’ HIV management goals of 95% awareness, 95% on treatment, and 95% [81]. A 3-year campaign mobilization on the role of clinical leaders in hypertension management at various health systems in the United States achieved the goal of 80% blood pressure control within seven months [97]. A scoping review of interventions to strengthen the health professionals’ leadership in SSA found that the opportunities for leadership development in SSA are scarce and those available are of poor quality and lacking a consistent evaluation framework [98]. There is a need for more studies on strengthening leadership in healthcare and exploring its impact on health outcomes, including hypertension in SSA.

As the burden of hypertension increases globally, the investment required of health systems to manage hypertension is rising. Hypertension is significantly associated with adverse outcomes of Coronavirus Disease 2019 (COVID-19), the ongoing pandemic ravaging the entire world’s health systems [99, 100]. Studies reported that hypertensive patients had almost three times the odds of mortality compared to people without hypertension [99]. The long-term post-COVID symptoms haunt the patients with hypertension who survive COVID-19 more than the general population [101]. For instance, patients with hypertension are twice as likely to have migraine-like headaches and 68% higher odds of poor sleep than people without hypertension [101]. Apart from the COVID-19 rationale, under the Sustainable Development Goal (SDG) 3 (Ensure healthy lives and promote well-being for all at all ages), target 3.4 aims to reduce by one-third the premature mortality from non-communicable diseases [102]. The management of hypertension, a significant risk factor for cardiovascular diseases, is a step closer to achieving target 3.4 of SDG 3.

Standard tools for exploring the health system determinants for specific diseases are scarce in the realm of hypertension care in Africa. There is a dire need for psychometric tools to explore health system readiness to provide quality hypertension care. The availability of those tools would open doors to researchers interested in hypertension care to gather quality data on health system determinants and render the evidence to policymakers to develop policies directed at shortfalls in care delivery to improve the health outcomes of patients with hypertension.

Future studies are recommended to explore the roles of leadership and governance and information systems at the health facilities levels as well as multi-level approaches (ie, individual, community, provider, health system) in the improvement of hypertension care [27, 72]. Additionally, while the current review explored studies conducted in SSA, most of the studies were conducted in three countries (Ethiopia, South Africa, and Ghana). Future studies should explore hypertension service readiness in other countries of SSA. The expansion of studies exploring health system factors of hypertension care could be a way to highlight the areas of need to advocate for support from local and international health agencies.

### Strengths and Limitations

The strengths of the current review are the holistic view of health system factors and interventions that affect the quality of hypertension service delivery, stringent critique of the quality of the studies, and the overall derivation conclusion against the WHO framework [25]. Nevertheless, the current review included only studies published in English and did not include the gray literature, which could consist of unpublished reports, and quality improvement projects on the health system interventions to improve hypertension service delivery. While the review attempted to isolate the effect of the individual health system factors, many system interventions were delivered in a bundled approach which rendered the isolation impossible.

## Conclusion

The literature exploring health system factors of hypertension outcomes in SSA is limited in volume and quality. The combination of multiple health system interventions was likely to result in better hypertension outcomes. More rigorously designed studies about the different aspects of the health systems and their impact on hypertension outcomes, especially the health information management and leadership and management are needed in SSA to inform effective hypertension control efforts.

## Supporting information

S-File 1. Search Terms

S-File 2. PRISMA 2009 Checklist

S-File 3. JBI Quality Assessment Results

S-File 4. SQUIRES

## Data Availability

The search terms used in the literature search have been provided in Supplementary File 1 and tables with the extracted data have been provided in the manuscript.

## List of abbreviations

CINAHL: Cumulated Index to Nursing and Allied Health Literature
JBI: Joanna Briggs Institute
HIV: Human Immunodeficiency Virus
LMIC: Low- and Middle-Income Countries
PRISMA: Preferred Reporting Items for Systematic Reviews and Meta-Analyses
SARA: Service Availability and Readiness Assessment
SDG: Sustainable Development Goals
COVID-19: Coronavirus Disease 2019
SQUIRES: Standards for Quality Improvement Reporting Excellence
SSA: Sub-Saharan Africa
WHO: World Health Organization

## Declarations

### Ethics approval and consent to participate

The current study did not involve human subject research, and the ethical approval or consent to participate is not applicable

### Consent for publication

All authors reviewed and approved the final manuscript for publication

### Competing interest

We have no conflict of interest to declare

### Funding

Not applicable

### Authors’ contributions

SB: Idea conception, literature search, title, and abstract review, full-text review, quality assessment, data extraction, analysis, and drafting of the manuscript

OO: Full-text review, quality assessment, manuscript drafting

FSS: Critical manuscript review, field expertise incorporation in the discussion of findings YCM: Critical manuscript review, language editing, supervision

KA: Full-text review, quality assessment, data extraction, analysis, and revision of the manuscript

CRH: Idea conception, design of literature search and inclusion and exclusion criteria, critical manuscript review, language editing, supervision

## Acknowledgments

We acknowledge the support of Stella Seal, the Library Informationist who helped in searching the databases, Dr. Sarah Szanton and Dr. William E. Rosa for the contribution of ideas in project ideation and discussion of findings, and Dr. Emmanuel Uwiringiyimana for the assistance in the full-text screening.

## Authors’ information

**Samuel Byiringiro**

Ph.D. Candidate at Johns Hopkins University School of Nursing

525 N Wolfe St, Baltimore, Maryland, 21205

United States

**Oluwabunmi Ogungbe**

Post-doctoral fellow at Johns Hopkins University School of Nursing

N Wolfe St, Baltimore, Maryland, 21205

United States

**Yvonne Commodore-Mensah**

Associate Professor at Johns Hopkins University School of Nursing

525 N Wolfe St, Baltimore, Maryland, 21205

United States

**Khadijat Adeleye**

Ph.D. Student at the University of Massachusetts

Elaine Marieb College of Nursing

Amherst, Massachusetts, 01002

United States of America

**Fred Stephen Sarfo**

Professor of Medicine at Kwame Nkrumah University of Science & Technology, Department of

Medicine

Accra Rd, Kumasi, Ghana

Clinician at Komfo Anokye Teaching Hospital

Okomfo Anokye Road, Kumasi, Ghana

**Cheryl R. Himmelfarb**

Sarah E. Allison Endowed Professor and Vice Dean for Research, Johns Hopkins University

School of Nursing, Joint Appointments in Schools of Medicine and Public Health

N Wolfe St, Baltimore, Maryland, 21205

United States

## Supporting information

S-File 1. Search terms

S-File 3. PRISMA 2009 checklist

S-File 3. JBI Quality Assessment Results

S-File 4. Standards for Quality Improvement Reporting Excellence

## Notes

### Competing Interest Statement

The authors have declared no competing interest.

### Funding Statement

None

### Author Declarations

We conducted online search for published literature

