## Supplementary material for "Health System Determinants of Hypertension Care and Outcomes in Sub-Saharan Africa: A Systematic Review": S-File 1. Search Terms

**Supplementary Table 1. Search terms as of June 27, 2022**

| **Database** | **Search Terms** | **Number of resulting Articles** |
| --- | --- | --- |
| PubMed | **(((("Health Facilities"[Mesh]) OR "Delivery of Health Care"[Mesh]) OR "Primary Health Care"[Mesh] OR "Health services" OR "Primary care" OR "Healthcare delivery" OR "Delivery of healthcare") AND (((("Africa South of the Sahara"[Mesh] OR “Central Africa” [tiab] OR Cameroon [tiab] OR “Central African Republic” [tiab] OR Chad [tiab] OR Congo [tiab] OR “Democratic Republic of the Congo” [tiab] OR “Equatorial Guinea” [tiab] OR Gabon [tiab] OR “Eastern Africa” [tiab] OR Burundi [tiab] OR Djibouti [tiab] OR Eritrea [tiab] OR Ethiopia [tiab] OR Kenya [tiab] OR Rwanda [tiab] OR Somalia [tiab] OR “South Sudan” [tiab] OR Sudan [tiab] OR Tanzania [tiab] OR Uganda [tiab] OR “Southern Africa” [tiab] OR Angola [tiab] OR Botswana [tiab] OR Lesotho [tiab] OR Malawi [tiab] OR Mozambique [tiab] OR Namibia [tiab] OR “South Africa” [tiab] OR Swaziland [tiab] OR Zambia [tiab] OR Zimbabwe [tiab] OR “Western Africa” [tiab] OR Benin [tiab] OR “Burkina Faso” [tiab] OR “Cape Verde” [tiab] OR “Cote d'Ivoire” [tiab] OR Gambia [tiab] OR Ghana [tiab] OR Guinea [tiab] OR “Guinea-Bissau” [tiab] OR Liberia [tiab] OR Mali [tiab] OR Mauritania [tiab] OR Niger [tiab] OR Nigeria [tiab] OR Senegal [tiab] OR “Sierra Leone” [tiab] OR Togo [tiab]))))) AND ("Hypertension"[Mesh] OR hypertens* [tiab] OR "high blood pressure" OR "raised blood pressure" OR "elevated blood pressure")** | 1840 |
| CINHAL | "((MH "Africa South of the Sahara+") OR ( ("Africa South of the Sahara" OR “Central Africa” OR Cameroon OR “Central African Republic” OR Chad OR Congo OR “Democratic Republic of the Congo” OR “Equatorial Guinea” OR Gabon OR “Eastern Africa” OR Burundi OR Djibouti OR Eritrea OR Ethiopia OR Kenya OR Rwanda OR Somalia OR “South Sudan” OR Sudan OR Tanzania OR Uganda OR “Southern Africa” OR Angola OR Botswana OR Lesotho OR Malawi OR Mozambique OR Namibia OR “South Africa” OR Swaziland OR Zambia OR Zimbabwe OR “Western Africa” OR Benin OR “Burkina Faso” OR “Cape Verde” OR “Cote d'Ivoire” OR Gambia OR Ghana OR Guinea OR “Guinea-Bissau” OR Liberia OR Mali OR Mauritania OR Niger OR Nigeria OR Senegal OR “Sierra Leone” OR Togo ) )) AND ((MH "Hypertension+") OR ( hypertens* OR ("blood pressure" N3 (elevat* OR high)) )) AND (( (MH "Health Facilities+") OR (MH "Health Services Accessibility+") OR (MH "Health Care Delivery+") ) OR ( (health N3 (facilit* OR service* OR accessibil*)) )) | 1135 |
| Embase | (('health care facilities and services'/exp OR 'health care facility'/de OR 'health care delivery'/de OR 'primary health care'/de) AND ('africa south of the sahara'/exp OR 'central africa' OR 'cameroon' OR 'central african republic' OR 'chad' OR 'congo' OR 'democratic republic of the congo' OR 'equatorial guinea' OR 'gabon' OR 'eastern africa' OR 'burundi' OR 'djibouti' OR 'eritrea' OR 'ethiopia' OR 'kenya' OR 'rwanda' OR somalia OR 'south sudan' OR sudan OR tanzania OR uganda OR 'southern africa' OR angola OR botswana OR lesotho OR malawi OR 'mozambique' OR 'namibia' OR 'south africa' OR 'swaziland' OR 'zambia' OR 'zimbabwe' OR 'western africa' OR 'benin' OR 'burkina faso' OR 'cape verde' OR 'côte d ivoire' OR 'ivory coast' OR 'gambia' OR 'ghana' OR 'guinea' OR 'guinea-bissau' OR 'liberia' OR 'mali' OR 'mauritania' OR 'niger' OR 'nigeria' OR 'senegal' OR 'sierra leone' OR 'togo') AND (('hypertension'/exp OR 'elevated blood pressure'/de OR 'abnormal blood pressure'/de OR hypertens*:ti,ab OR 'high blood pressure') NOT 'pregnancy' NOT 'eclampsia' NOT 'preeclampsia')) AND **'article'**/it AND (**2010**:py OR **2011**:py OR **2012**:py OR **2013**:py OR **2014**:py OR **2015**:py OR **2016**:py OR **2017**:py OR **2018**:py OR **2019**:py OR **2020**:py OR **2021**:py OR **2022**:py) | 4519 |
