## Supplementary material for "Health System Determinants of Hypertension Care and Outcomes in Sub-Saharan Africa: A Systematic Review": S-File 3. JBI Quality Assessment Results

**Supplementary File 2. JBI Quality Assessment Tools**

Risk of bias for randomized controlled trials

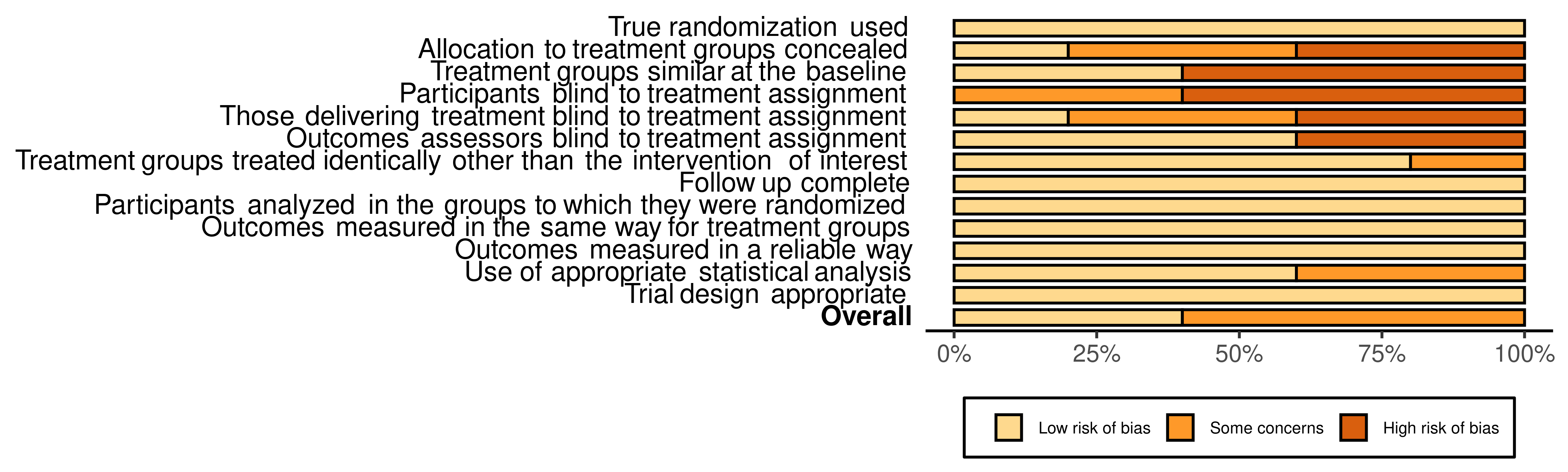

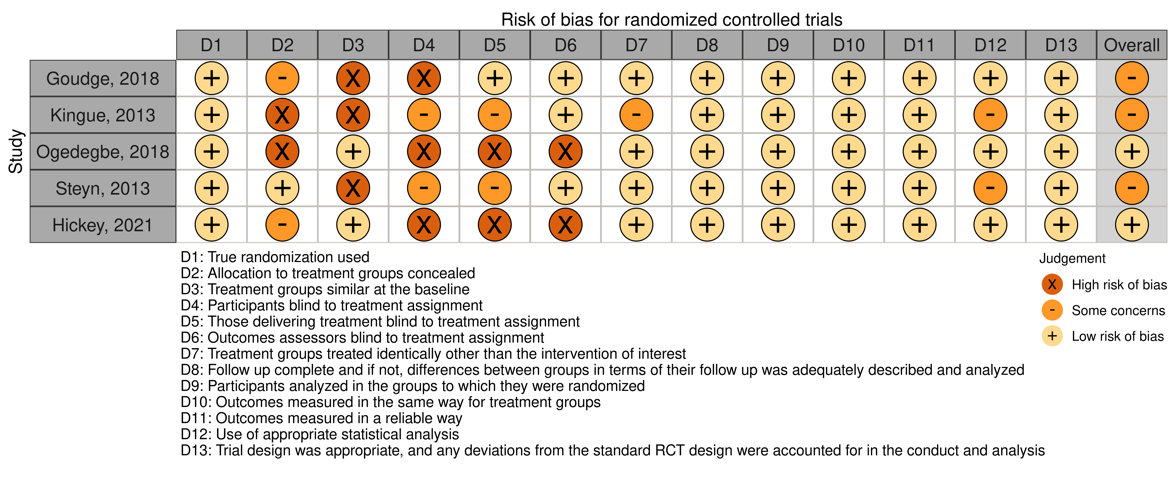

Risk of bias for quasi-experimental studies
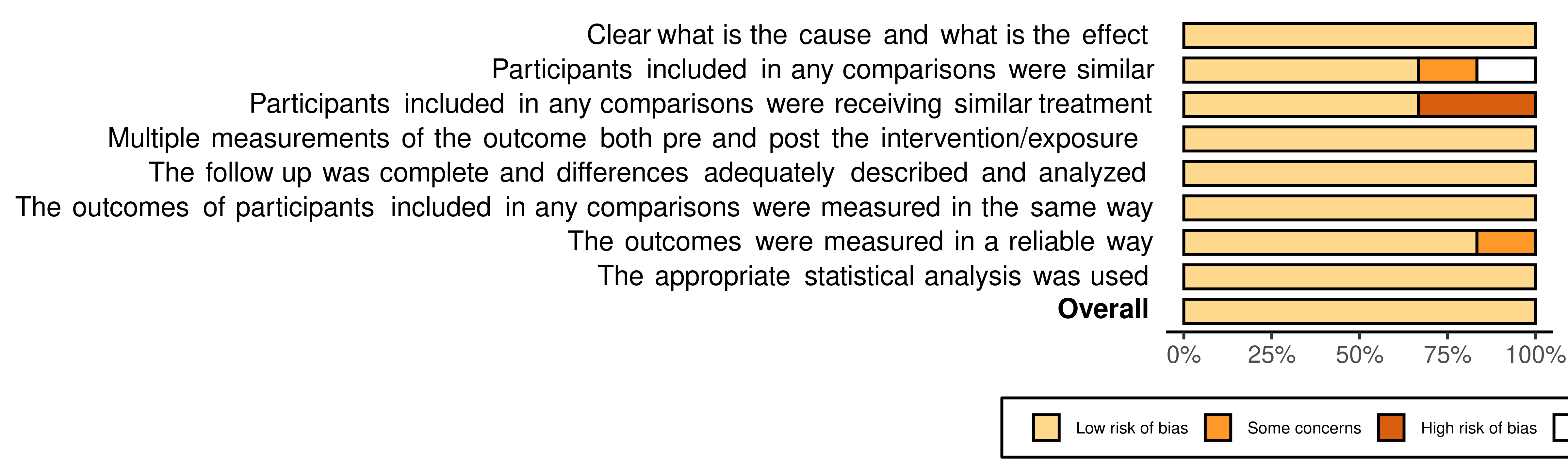

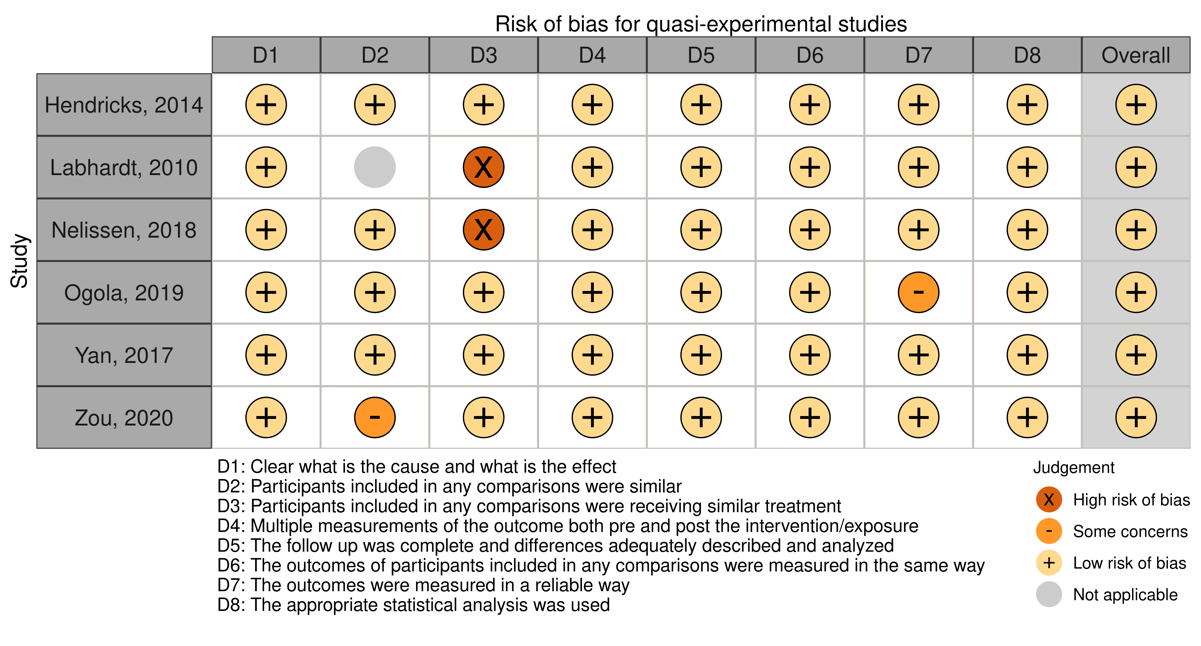

Risk of bias assessment for cohort studies

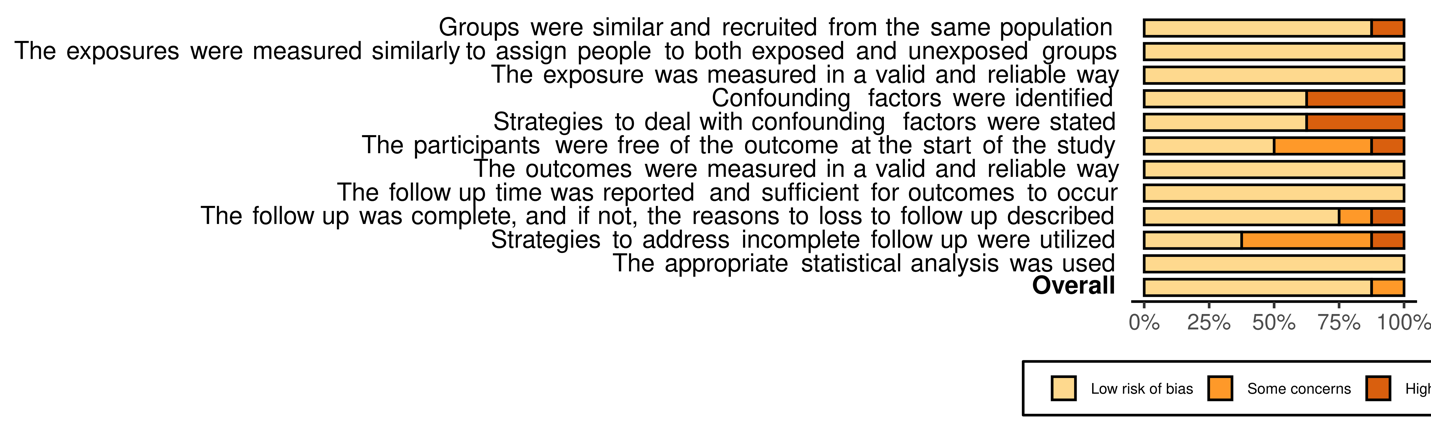

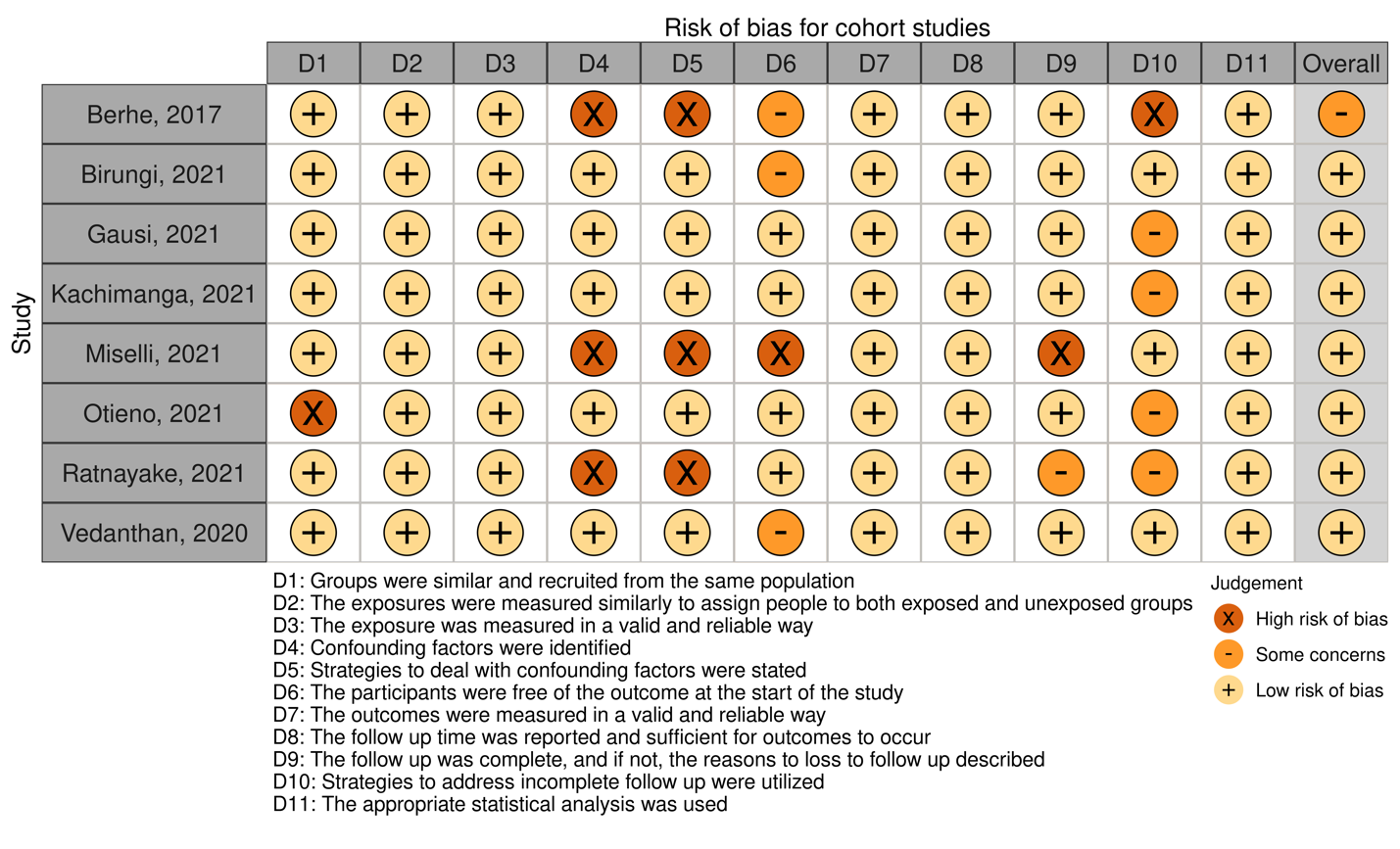

Risk of bias for cross-sectional studies

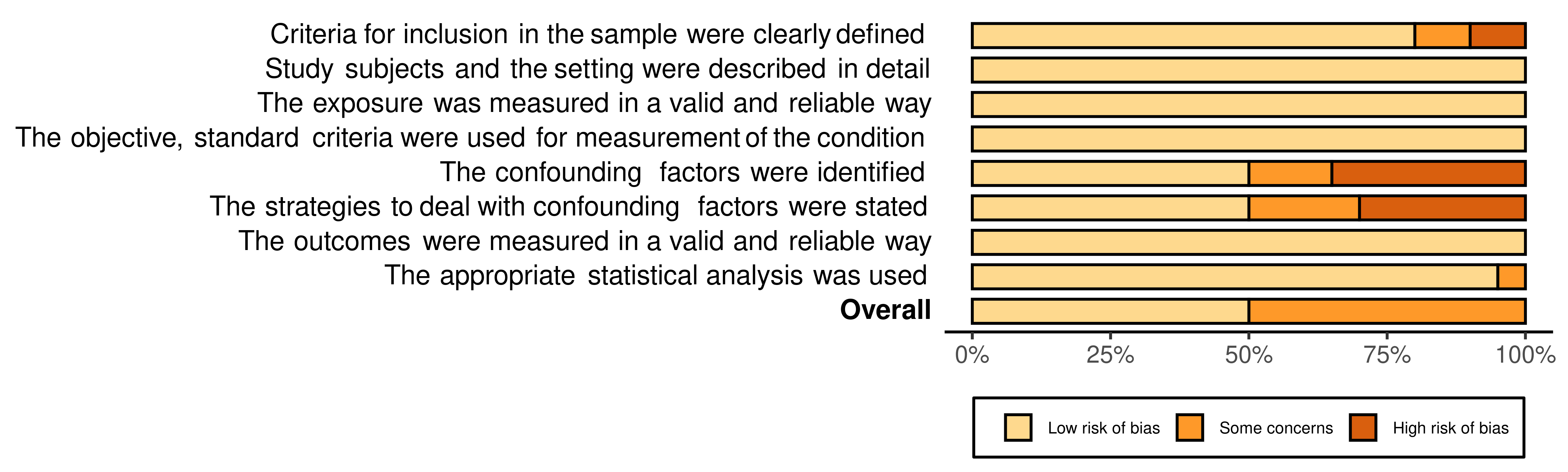

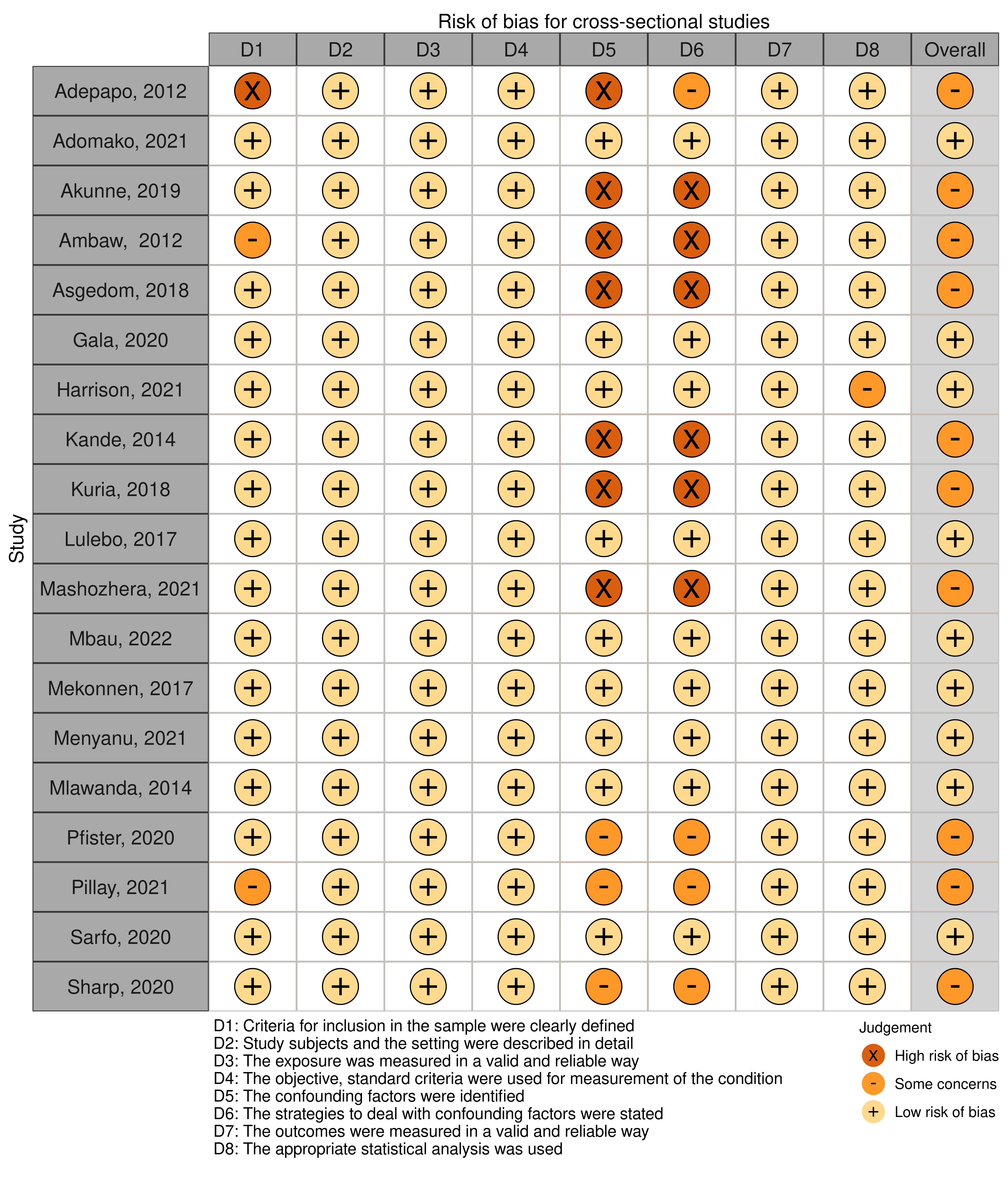
