## Supplementary material for "Health System Determinants of Hypertension Care and Outcomes in Sub-Saharan Africa: A Systematic Review": S-File 4. SQUIRES

**Supplementary File 3. Standards for Quality Improvement Reporting Excellence (SQUIRE 2.0)** [1]**: Assessment of the study:** Improving the quality of care for patients with hypertension in Moshupa District, Botswana: Quality improvement cycle [2].

| **Text Section and Item Name** | **Section or Item Description** | |  |
| --- | --- | --- | --- |
| **Notes to authors** | - The SQUIRE guidelines provide a framework for reporting new knowledge about how to improve healthcare      - The SQUIRE guidelines are intended for reports that describe system level work to improve the quality, safety, and value of healthcare, and used methods to establish that observed outcomes were due to the intervention(s).      - A range of approaches exists for improving healthcare. SQUIRE may be adapted for reporting any of these.      - Authors should consider every SQUIRE item, but it may be inappropriate or unnecessary to include every SQUIRE element in a particular manuscript.      - The SQUIRE Glossary contains definitions of many of the key words in SQUIRE.      - The Explanation and Elaboration document provides specific examples of well-written SQUIRE items, and an in-depth explanation of each item.      - Please cite SQUIRE when it is used to write a manuscript. | | **As you review the manuscript, place a checkmark in this column for each**  **SQUIRE item that is appropriately**  **addressed in the manuscript.**  **Remember that not every item is**  **necessary in every manuscript.** |
| **Title and Abstract** |  | |  |
| **1. Title** | Indicate that the manuscript concerns an initiative to improve healthcare (broadly defined to include the quality, safety, effectiveness, patient-centeredness, timeliness, cost, efficiency, and equity of healthcare) | | YES |
| **2. Abstract** | a. Provide adequate information to aid in searching and indexing b. Summarize all key information from various sections of the text using the abstract format of the intended publication or a structured summary such as: background, local problem, methods, interventions, results, conclusions | | YES |
| **Introduction** | *Why did you start?* | |  |
| **3. Problem Description** | Nature and significance of the local problem | | YES |
| **4. Available knowledge** | Summary of what is currently known about the problem, including relevant previous studies | | YES |
| **5. Rationale** | Informal or formal frameworks, models, concepts, and/or theories used to explain the problem, any reasons or assumptions that were used to develop the intervention(s), and reasons why the intervention(s) was expected to work | | YES |
| **6. Specific aims** | Purpose of the project and of this report | | YES |
| **Methods** | *What did you do?* | |  |
| **7. Context** | Contextual elements considered important at the outset of introducing the intervention(s) | | YES |
| **8. Intervention(s)** | a. | Description of the intervention(s) in sufficient detail that others could reproduce it | Intervention (Not clearly described)  Team (YES) |
|  | b. | Specifics of the team involved in the work |  |
| **9. Study of the**  **Intervention(s)** | a.  b. | Approach chosen for assessing the impact of the intervention(s)  Approach used to establish whether the observed outcomes were due to the intervention(s) | YES |
| **10. Measures** | a.  b. | Measures chosen for studying processes and outcomes of the intervention(s), including rationale for choosing them, their operational definitions, and their validity and reliability  Description of the approach to the ongoing assessment of contextual elements that contributed to the success, failure, efficiency, and cost | YES |
|  | c. | Methods employed for assessing completeness and accuracy of data |  |
| **11. Analysis** | a.  b. | Qualitative and quantitative methods used to draw inferences from the data  Methods for understanding variation within the data, including the effects of time as a variable | YES |
| **12. Ethical**  **Considerations** | Ethical aspects of implementing and studying the intervention(s) and how they were addressed, including, but not limited to, formal ethics review and potential conflict(s) of interest | | YES |

| **Results** | *What did you find?* |  |
| --- | --- | --- |
| **13. Results** | a. Initial steps of the intervention(s) and their evolution over time (*e.g.*, time-line diagram, flow chart, or table), including modifications made to the intervention during the project b. Details of the process measures and outcome c. Contextual elements that interacted with the intervention(s) d. Observed associations between outcomes, interventions, and relevant contextual elements e. Unintended consequences such as unexpected benefits, problems, failures, or costs associated with the intervention(s). f. Details about missing data | YES |
| **Discussion** | *What does it mean?* |  |
| **14. Summary** | a. Key findings, including relevance to the rationale and specific aims b. Particular strengths of the project | YES |
| **15. Interpretation** | a. Nature of the association between the intervention(s) and the outcomes b. Comparison of results with findings from other publications c. Impact of the project on people and systems d. Reasons for any differences between observed and anticipated outcomes, including the influence of context e. Costs and strategic trade-offs, including opportunity costs | YES |
| **16. Limitations** | a. Limits to the generalizability of the work b. Factors that might have limited internal validity such as confounding, bias, or imprecision in the design, methods, measurement, or analysis c. Efforts made to minimize and adjust for limitations | YES |
| **17. Conclusions** | a. Usefulness of the work b. Sustainability c. Potential for spread to other contexts d. Implications for practice and for further study in the field e. Suggested next steps | YES |
| **Other information** |  |  |
| **18. Funding** | Sources of funding that supported this work. Role, if any, of the funding organization in the design, implementation, interpretation, and reporting | YES/NO (Declared lack of any conflict of interest but did not specify source of funding) |
